# Performance of a Chinese Cognitive Decline Risk Model in a Japanese Cohort: A Validation Study

**DOI:** 10.64898/2025.12.25.25342996

**Authors:** Lihui Tu, May Kristine Jonson Carlon, Yoshinori Nanjo, Dongmei Gu, Yasuo Kuniyoshi

## Abstract

**Objectives:** To develop a simple risk prediction model for cognitive decline in a Chinese older adult cohort, and to evaluate its performance and transportability through temporal validation and external validation in a Japanese older adult cohort.

**Methods:** The prediction model was developed using a derivation cohort of 5,985 cognitively normal older adults from the China Health and Retirement Longitudinal Study (CHARLS, 2011-2015). A comparison of seven machine learning algorithms was conducted, and the standard Cox Proportional Hazards (CoxPH) model was selected based on its optimal balance of performance and parsimony. The final model was then validated on a temporal cohort (CHARLS 2015-2018, n=1,333) and an external cohort (Japanese Study of Aging and Retirement [JSTAR] 2007-2009, n=2,798). A comprehensive preprocessing pipeline, including Iterative Imputation for high-missingness predictor variables and One-Hot Encoding for categorical variables, was developed on the training data and applied to all cohorts. Model performance was assessed via discrimination, calibration, risk stratification and clinical utility.

**Results:** In temporal validation, the model demonstrated strong performance with an AUC of 0.72 and reliable calibration (Slope = 1.02). In the external JSTAR cohort, the model maintained high discriminative power (AUC = 0.68), which was even superior to the development set (AUC = 0.62). However, a notable calibration shift was observed (Slope = 1.54), indicating a systematic underestimation of absolute risk in the low-prevalence Japanese population. While decision curve analysis (DCA) showed substantial net benefit in the temporal cohort, its utility in the external cohort was most effective within a narrow threshold range near the population prevalence. Sensitivity analyses confirmed that the model’s risk-ranking ability remained robust across 2-year and 4-year horizons.

**Conclusion:** Our 6-predictor model shows robust risk-ranking consistency across cohorts, but absolute risk estimates are sensitive to population and temporal differences. While effective for identifying high-risk individuals, local recalibration is essential for accurate clinical prognosis in new settings.

## 1. Introduction

Dementia, a serious stage of cognitive impairment, is recognized for imposing a heavy burden on both individuals and society (1,2). As current treatments are often initiated late and demonstrate limited efficacy (3,4), timely intervention necessitates earlier identification. This is of particular interest at the earliest possible stage: cognitive decline per se. While a small proportion of individuals will experience cognitive decline, (5,6) numerous associated factors have been thoroughly sought. This indicates that predicting the onset of cognitive impairment from a healthy state may be accessible.

In the context of big data and machine learning (ML), several previous studies have focused on factors associated with cognitive decline (7–11). Overall, the predictive accuracy of existing models varies widely (area under curve [AUC] 0.6 ∼ 0.9) depending on the cohort population, feature options, and outcome definitions (8,12). While most models are able to discriminate cognitive decline, a more glaring issue, however, is the lack of external validation, particularly cross-national validation (7,13). Overreliance on internal validation where test and training datasets are highly similar can lead to over-optimistic results (14). This limits the model’s generalizability and clinical utility.

Rigorous external validation serves several crucial purposes (15–18). First, it helps minimize overfitting and determine the robustness of the model. Second, it allows for an investigation of associated factors in a broader population context, which helps identify universal determinants across diverse populations. This is essential for achieving the generalizability and applicability required for clinical practice.

The major objective of this study was to develop a prediction model for cognitive decline in a Chinese cohort using an ML approach and validate this model in a Japanese cohort. As two major East Asian populations, China and Japan share certain similarities but also possess distinct social and healthcare contexts. This cross-national validation, therefore, provides a robust test of the model’s transportability. To assess the model’s robustness over time before external validation, we also conducted a temporal validation using a later wave of the same Chinese cohort.

## 2. Methods

### 2.1 Study Design and Cohorts

#### 2.1.1 Derivation cohort (CHARLS 2011-2015)

Data for model development were drawn from the China Health and Retirement Longitudinal Study (CHARLS). CHARLS is an ongoing, nationally representative, and prospective cohort study of community-dwelling Chinese residents aged 45 years and older. Baseline examinations were conducted in 2011 (N = 17,708), with follow-up surveys conducted every two years. The survey provides information on demographics, health status, behavior, and cognition (19).

For this study, the derivation cohort was defined as participants from the CHARLS 2011-2015 dataset, encompassing a 4-year follow-up period. Inclusion criteria were defined as: (1) aged 50 years or older at baseline (2011); (2) were cognitively healthy at baseline (defined in Outcome Assessment); and (3) had follow-up cognitive data available in 2013 (wave2) and 2015 (wave 3).

#### 2.1.2 Validation Cohorts

Two independent cohorts were used for temporal and external validation, respectively. For temporal validation, we used a subsequent, non-overlapping CHARLS cohort (2015-2018). Participants from the 2015 wave who met the same inclusion criteria were included and followed up for 3 years (to 2018). This approach was used to examine model performance in the same underlying population but at a different time point. Temporal validation helps ensure that the model is not overfitted to time-specific patterns, thus providing a stronger basis for assessing its external generalizability.

For external validation, we used data from the Japanese Study of Aging and Retirement (JSTAR). JSTAR is a prospective cohort study initiated in 2007, following participants aged 50 and older, and like CHARLS, was patterned after the US Health and Retirement Study (20). Due to cognitive measures only being available in the first two waves, the JSTAR 2007-2009 cohort was included as the final external validation set. This external validation provides an opportunity to assess the model’s generalizability across distinct East Asian populations and to examine its robustness in a different social and healthcare context.

### 2.2 Outcome Assessment

Cognitive function in CHARLS was assessed using measures of memory (word recall) and orientation. Consistent with previous literature indicating memory impairment as a primary indicator of Alzheimer’s dementia (21), cognitive decline was defined based solely on the memory score. A cutoff value was developed within the derivation cohort using an education-stratified threshold (No formal schooling, Primary/Middle, High School and above) of mean minus one standard deviation (SD). Individuals were defined as cognitively healthy at baseline if their memory score was at or above this cutoff. Incident cognitive decline (the primary outcome, event=1) was defined as a participant’s memory score dropping below this established, education-specific cutoff at the follow-up wave.

### 2.3 Predictor Assessment (Baseline Features)

We aimed to build a community-based prediction model that could be easily implemented for mass screening. Therefore, baseline features selected for model prediction were focused on common and easily accessible factors across cohorts. Predictors were selected covering several categories: (1) sociodemographic factors: age, sex, education, and marital status; (2) health status and cognition: body mass index, waist circumference, grip strength, chronic vascular disease status, subjective memory problem, sleep quality, and baseline memory and orientation scores; (3) physical function: activities of daily living impairment; and (4) lifestyle and psychosocial factors: smoke status, alcohol intake, life satisfaction, and depressive symptoms. Following the model development and selection process detailed below, a final, parsimonious set of six predictors was identified.

### 2.4 Statistical Analysis

#### 2.4.1 Data Preprocessing

A comprehensive preprocessing pipeline was developed using scikit-learn to ensure consistent data transformation across all datasets and prevent data leakage during cross-validation (CV). For continuous variables, two strategies were used based on missingness observed in the training data. For grip strength (10-20% missing), missing values were imputed using IterativeImputer (MICE) with BayesianRidge regression as the proportion of missingness may suggest a potential deviation from randomness. The other continuous variable, orientation score (<5% missing), was imputed using SimpleImputer (strategy=’median’) to not introduce unnecessary complexity brought about by MICE on such a small number of missing values. All continuous variables were subsequently scaled using StandardScaler. Categorical variables were converted to dummy variables using OneHotEncoder (drop=’first’).

#### 2.4.2 Model Development and Selection

Numerous ML methods have been investigated in the past (10,15) including unsupervised and supervised approaches. For our approach, survival models were selected as the primary framework. (22) This approach was chosen because it appropriately handles censored data and utilizes the full time-to-event information, which is more robust than simple logistic regression at a fixed time point. Therefore, the model inherently possesses the flexibility to provide risk predictions at any specific time interval (e.g., 2-year or 3-year), even when trained on a cohort with a median follow-up of 4 years. The derivation cohort was first split into a training set (80%) and a final hold-out test set (20%). To select the optimal model, a 10-fold stratified cross-validation (CV) was performed on the 80% training set.

Specifically, seven distinct ML algorithms were examined, incorporating the preprocessing pipeline. These included linear models: CoxPH Model without elastic net penalty, ElasticNet-CoxPH; and tree-based ensembles: Gradient Boosting Survival Analysis (GBSA), Random Survival Forest (RSF), ExtraTrees Regressor (EST); and XGBoost-Cox, and LightGBM-Reg.

#### 2.4.3 Feature Selection and Parsimony

To select the most relevant and parsimonious set of predictors from the full candidate list, a multi-step feature reduction process was employed. First, all candidate predictors were ranked using permutation feature importance derived from the RSF model, which was fit on the 80% training set. This was done as tree-based methods can capture nonlinear and interaction effects among predictors. Second, a stepwise selection process was conducted based on this ranking. Starting with the top-ranked features, predictors were iteratively added to the base CoxPH model (the chosen architecture).

Finally, the model’s performance (assessed by concordance index [C-index] and integrated Brier score [IBS] in CV) was balanced against model complexity (number of variables). This process identified a final, parsimonious set of six predictors (age, sex, education, grip strength, memory, and orientation) that provided optimal predictive performance while maintaining simplicity for potential clinical implementation.

#### 2.4.4 Model Selection and Ranking

Within each fold of the CV, model performance was assessed across multiple domains. First by discrimination which is measured by Harrell’s C-index (C-index) and time-dependent AUC (CD-AUC) at 4 years. Second is by calibration which is measured by the IBS and Root Mean Squared Error (RMSE) over the follow-up period. Models were ranked on each metric, and a ‘Mean Rank’ was used to identify the most balanced and high-performing model architecture.

#### 2.4.5 Final Model Stratification and Evaluation

Following the selection of the optimal model architecture, we performed a comprehensive evaluation of its clinical utility. First, for risk stratification, we employed a Bayesian Gaussian Mixture Model (BGMM) to categorize the predicted risk scores (LogHR) into Low, Medium, and High risk tiers. Unlike fixed-threshold methods, BGMM is an unsupervised approach that identifies latent risk phenotypes by fitting multiple Gaussian distributions to the score density. Second, the discriminative power and calibration of these categories were assessed using Kaplan-Meier survival analysis with log-rank tests. To ensure the stability of the stratification, this process was mirrored in both the 80% cross-validation and the 20% internal testing sets.

#### 2.4.6 Temporal and External Validation

The champion model architecture, selected from the 10-fold CV, was then re-trained on the entire 80% training set (using the corresponding preprocessing pipeline). This single, final model was used for all subsequent evaluations. Performance was first assessed on the 20% internal hold-out test set. Subsequently, the model’s performance was validated on the two independent cohorts.

Performance in these cohorts was investigated using the same metrics, supplemented by visual analysis of discrimination with Receiver Operating Characteristic (ROC) curves at the respective follow-up end points and calibration with calibration plots (Observed vs. Predicted risk) at the same time points. Risk stratification was also evaluated by comparing cumulative incidence. An optimal cutoff for the model’s predicted risk score was determined in the development cohort using maximally selected rank statistics (MaxStat). This pre-defined cutoff was then applied to the validation cohorts to stratify participants into different groups. The cumulative incidence of cognitive decline was then compared between these groups. Decision Curve Analysis (DCA) to quantify the net benefit in terms of clinical utility. To explore the potential influence of cohort heterogeneity, predicted risk distributions (“case-mix”) were compared across validation datasets using histograms and the Kolmogorov–Smirnov statistic.

### 2.5 Sensitivity Analyses

Given the discrepancy in follow-up durations between the training cohort (4 years) and the external validation cohorts (2–3 years), we conducted a sensitivity analysis within the training set to evaluate the model’s performance at different time horizons.

Two other sensitivity analyses were also conducted on the JSTAR external validation cohort to assess the robustness of our findings. First, to assess the impact of the imputation strategy, a complete-case analysis was performed. The final model pipeline (including preprocessing) was re-trained only on the subset of the CHARLS development cohort with no missing data in the 6 key predictors, and subsequently validated only on the subset of the JSTAR cohort who also had complete data.

Second, to assess model transportability and the impact of distributional shift, we tested the effect of "re-calibration" of the preprocessor. The original CoxPH coefficients (trained on CHARLS) were retained, but the preprocessing pipeline was re-fit on the JSTAR cohort (i.e., using JSTAR’s own median, imputation models, and scaling parameters) before predicting risk.

The performance metrics from both sensitivity analyses were then compared to those of the primary validation. All analyses were conducted using Python (v3.10) with the scikit-survival (v0.21), lifelines (v0.27), and scikit-learn (v1.1) libraries. A two-sided p<0.05 was considered statistically significant.

## 3. Results

### 3.1 Participant Characteristics

The participant selection flowchart is presented in **Figure 1**. After applying the inclusion and exclusion criteria, the derivation cohort (CHARLS 2011-2015) included 5,985 participants, with 1,333 and 2,798 participants enrolled in the temporal (CHARLS 2015-2018) and external (JSTAR 2007-2009) validation cohorts, respectively.

**Figure 1.**
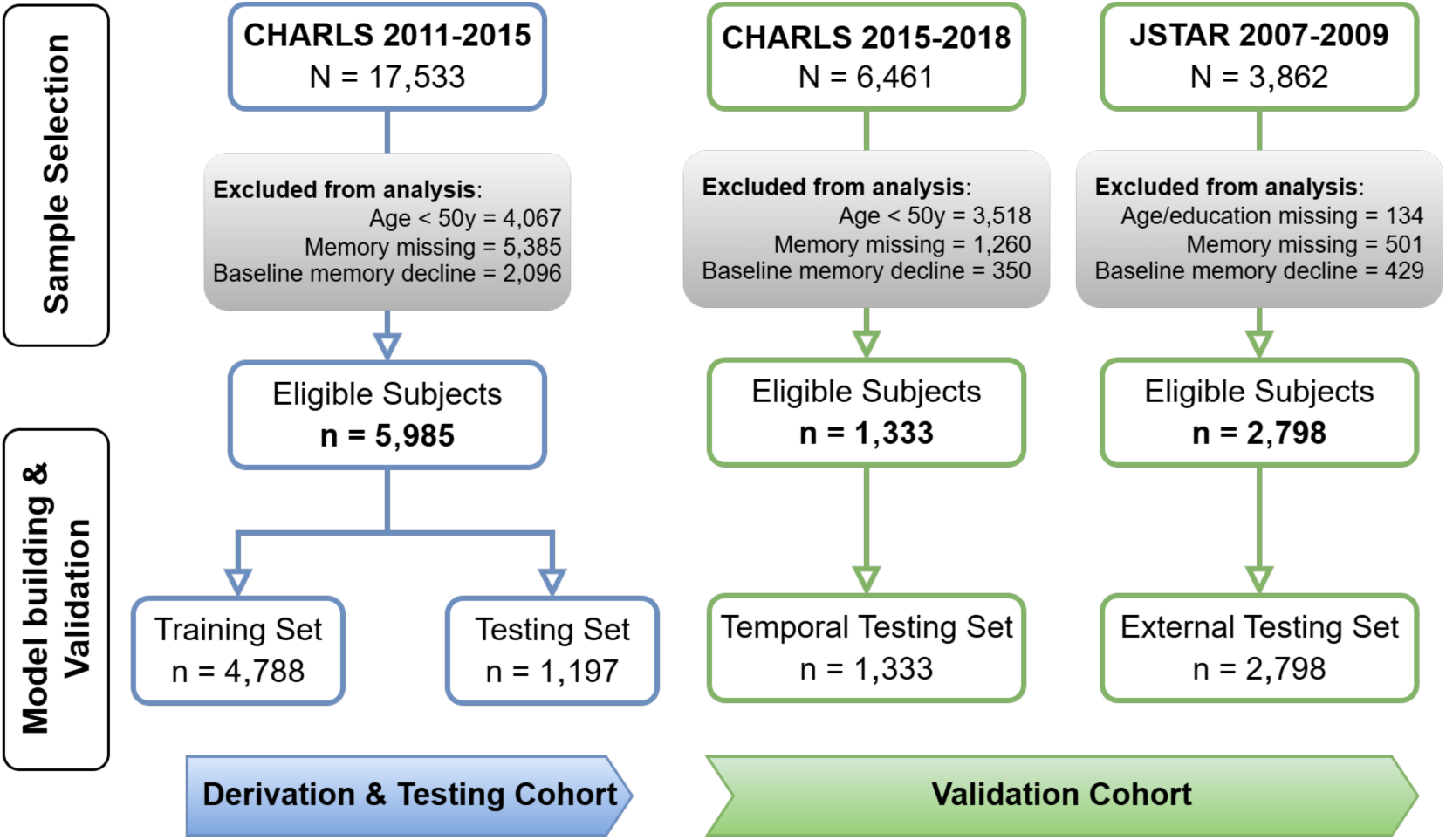
Cohort and subject selection. This flow diagram shows subject inclusion and exclusion in each cohort, as well as the data set partition for training, temporal and external validation.

The baseline characteristics are detailed in **Table 1**. Compared to the derivation cohort, both validation cohorts exhibited certain heterogeneities. Participants in the temporal cohort were slightly younger and possessed higher baseline cognitive scores, indicating a relatively healthier profile at baseline. Notably, participants in the external JSTAR cohort were older, had a higher proportion of females, and displayed substantially higher education levels, suggesting significant cross-population heterogeneity that must be considered when interpreting model generalizability.

**Table 1.** Baseline characteristics by cohort.

| Variable | Training Cohort<br>CHARLS<br>(n=5,985) | Validation Cohort |  |  |  |
| --- | --- | --- | --- | --- | --- |
|  |  | CHARLS<br>(n=1,333) | *p* * | JSTAR<br>(n=2,798) | *p* ** |
| Sociodemographic factors |  |  |  |  |  |
| Age and group | 59.0(55.0,65.0) | 58.0(52.0,64.0) | <0.001 | 62.0(57.0,68.0) | <0.001 |
| <65 | 4,457 (74.5) | 1,008 (75.6) |  | 1,693 (60.5) |  |
| 65–74 | 1,288 (21.5) | 264 (19.8) | 0.279 | 1,062 (38.0) | <0.001 |
| ≥75 | 240 (4.0) | 61 (4.6) |  | 43 (1.5) |  |
| Female | 3,008 (50.3) | 637 (47.8) | 0.108 | 1,478 (52.8) | 0.026 |
| Education group |  |  |  |  |  |
| No formal schooling | 1,678 (28.0) | 236 (17.7) | <0.001 | 0 (0.0) | <0.001 |
| Primary/Middle | 3,585 (59.9) | 859 (64.5) |  | 810 (28.9) |  |
| High School and above | 722 (12.1) | 238 (17.9) |  | 1,988 (71.1) |  |
| Physical function |  |  |  |  |  |
| Grip strength<sup>§</sup> | 32.0 (25.5,40.0) | 31.5 (25.0,40.0) | 0.983 | 28.0 (22.0,36.0) | <0.001 |
| Cognitive performance |  |  |  |  |  |
| Memory score | 8.0(6.0,10.0) | 8.0 (6.0,10.0) | <0.001 | 10.0 (9.0,12.0) | <0.001 |
| Orient score<sup>§§</sup> | 3.0 (3.0,4.0) | 4.0 (3.0,4.0) | <0.001 | 6.0 (6.0,6.0) | <0.001 |

Data are presented as median (IQR) or n (%).
Mann–Whitney U test for continuous variables, and chi-square for categorical variables.
*p*\* Validation CHARLS vs. training cohort; *p*\*\* Validation JSTAR vs. training cohort.
<sup>§</sup> Missing data: 713(11.9%), 267(20.0%), and 56(2.0%), respectively.
<sup>§§</sup> Missing data: 225(3.8%), 37(2.8%), and 0, respectively.

### 3.2 Final Model Parameters and Feature Selection

Performance metrics for the candidate ML models are summarized in **Table 2**. The standard CoxPH model was identified as the optimal approach, achieving a C-index of 0.627 and a CD-AUC of 0.656. The initial ranking by RSF permutation importance (**Supplementary Figure 1**) identified the top predictors. Following the multi-step process of balancing performance against parsimony, a final six-predictor model was selected.

**Table 2.** Performance metrics of machine learning models during cross-validation.

| Model | C-index | CD-AUC | IBS | RMSE | Mean Rank |
| --- | --- | --- | --- | --- | --- |
| **Linear models** |  |  |  |  |  |
| CoxPH Model without elastic net penalty | **0.627 (0.597–0.657)** | **0.656 (0.632–0.680)** | **0.120 (0.115–0.124)** | 0.016 (0.013–0.020) | **1.5** |
| ElasticNet-CoxPH | 0.584 (0.566–0.602) | 0.609 (0.597–0.620) | 0.125 (0.119–0.130) | **0.014 (0.010–0.018)** | 2.75 |
| **Ensemble models** |  |  |  |  |  |
| Gradient Boosting Survival Analysis (GBSA) | 0.613 (0.581–0.645) | 0.648 (0.623–0.672) | 0.137 (0.135–0.140) | 0.127 (0.119–0.135) | 3.25 |
| Random Survival Forest (RSF) | 0.578 (0.556–0.601) | 0.608 (0.587–0.630) | 0.129 (0.122–0.135) | 0.015 (0.011–0.019) | 3.75 |
| eXtreme Gradient Boosting Cox (XGBoost-Cox) | 0.608 (0.578–0.637) | 0.641 (0.617–0.665) | 0.241 (0.232–0.250) | 0.190 (0.181–0.199) | 5 |
| Light Gradient Boosting Machine-Reg (LightGBM-Reg) | 0.535 (0.502–0.567) | 0.574 (0.543–0.605) | 0.139 (0.135–0.142) | 0.116 (0.109–0.124) | 5.5 |
| Extra Survival Trees (EST) | 0.547 (0.525–0.569) | 0.562 (0.542–0.582) | 0.152 (0.147–0.156) | 0.131 (0.123–0.139) | 6.25 |

Values represent the mean and 95% confidence interval.
C-index, concordance index based on inverse probability of censoring weights; CD-AUC, cumulative dynamic area under the curve; IBS, integrated Brier score; RMSE, root mean square error.

The final model parameters are presented in **Table 3**. Consistent with the consensus that demographic and functional factors drive cognitive health, older age, male sex, and lower education were associated with an increased hazard of decline. Conversely, better baseline cognitive function and stronger grip strength were found to be protective, reinforcing their roles as key indicators of cognitive reserve.

**Table 3.**
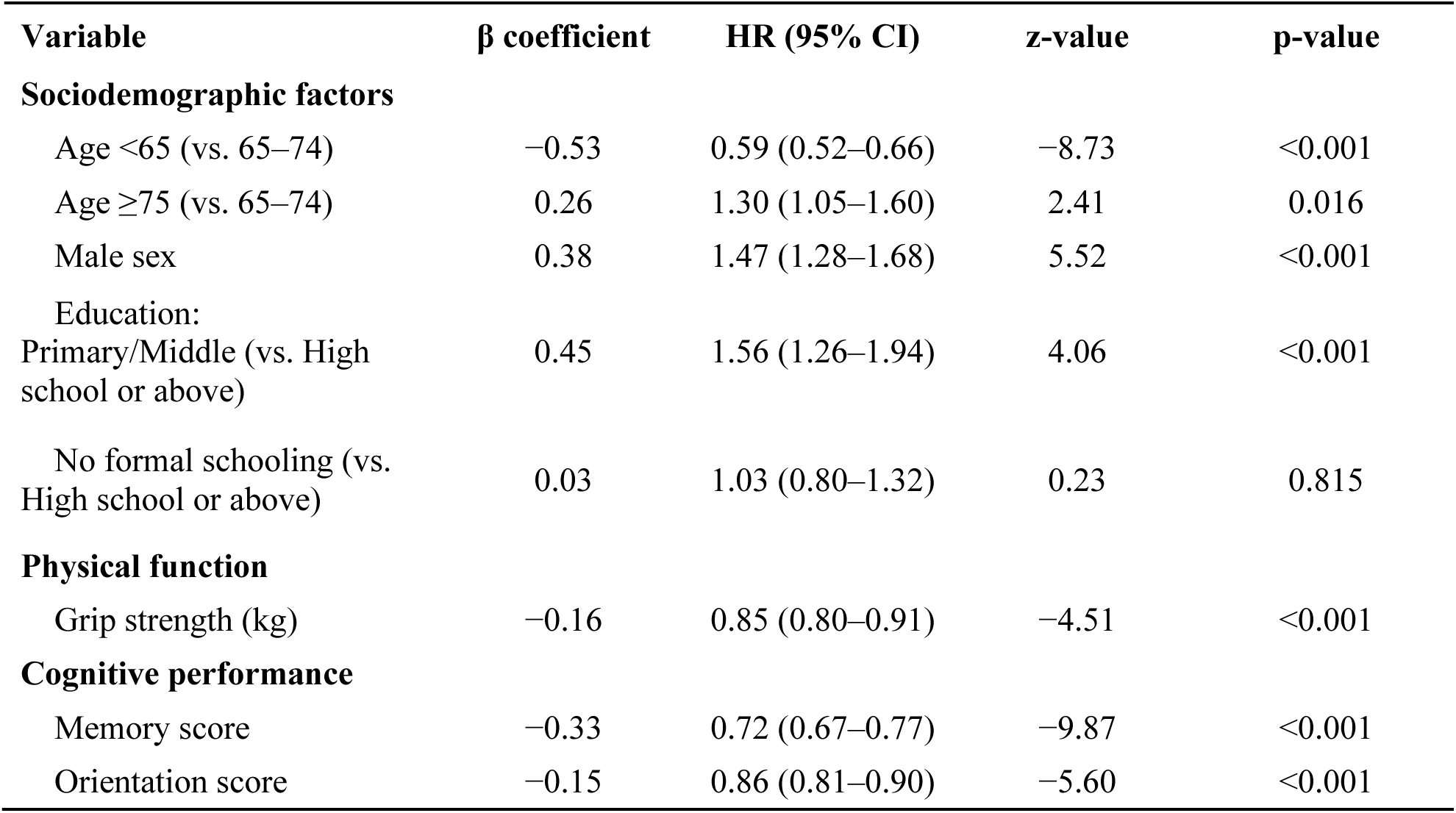
Key predictors of cognitive decline in the development cohort (CoxPH Model)

### 3.3 Predictive Performance and Generalizability

#### 3.3.1 Discriminative Ability Across Cohorts

The 6-predictor model demonstrated stable discrimination across the hold-out test set and independent cohorts (**Supplementary Table**). Unexpectedly, a discrepancy was observed between internal and external evaluations: while the internal test set yielded a modest C-index (0.609), the temporal (0.722) and external (0.682) validation cohorts showed substantially higher performance. This robust performance is further supported by the corresponding ROC curves (**Figure 2A**). We speculate that this difference might partly reflect sampling variability in the smaller internal set and the greater heterogeneity in validation cohorts, which potentially reveals a broader discriminative range for the model.

**Figure 2.**
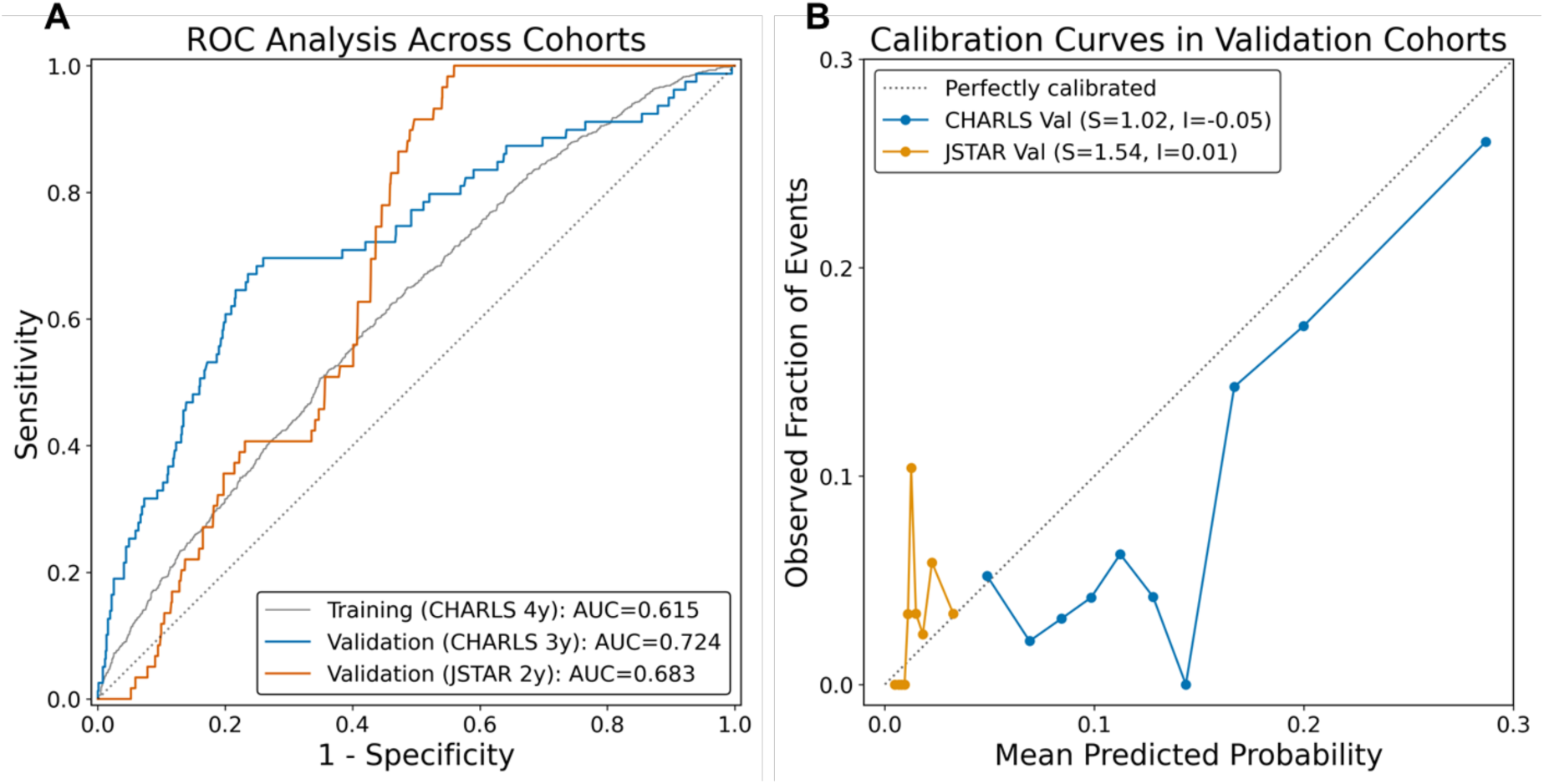
Discriminative ability and calibration across cohorts. **(A)** Receiver Operating Characteristic (ROC) curves across the three study cohorts. The model maintained robust discrimination with AUCs of 0.615 for the training set (CHARLS 4y), 0.724 for the temporal validation (CHARLS 3y), and 0.683 for the external validation (JSTAR 2y). **(B)** Calibration plots comparing predicted risk with observed cognitive decline incidence. The model demonstrated excellent calibration in the temporal validation cohort (Slope = 1.02, Intercept = -0.05). In the external JSTAR cohort, the model showed moderate calibration (Slope = 1.54, Intercept = 0.01), with the slope suggesting a tendency to underestimate risk in this extremely low-incidence population.

Case-mix comparison (**Supplementary Figure 2**) further clarified these results. Predicted risks in CHARLS were broadly distributed, consistent with its moderate event rate (5.85%). In contrast, the JSTAR cohort displayed an extremely low-risk profile, with nearly all predicted probabilities concentrated below 0.02, reflecting its lower observed 2-year incidence (0.68%).

#### 3.3.2 Model Calibration and Potential Clinical Utility

The model demonstrated excellent calibration in the temporal validation cohort (Slope = 1.02, Intercept = -0.05, **Figure 2B**). In the JSTAR cohort, a moderate calibration was observed (Slope = 1.54, Intercept = 0.01, **Figure 2B**). The higher slope suggests a tendency to underestimate risk in this extremely low-incidence population, which might be considered a conservative estimation bias.

Regarding clinical utility, DCA demonstrated a clear net benefit in the temporal cohort within a threshold range of 0.05 to 0.12 (**Supplementary Figure 3A**). While the net benefit was more marginal in the JSTAR cohort (**Supplementary Figure 3B**), the model still demonstrated positive utility at lower threshold ranges, suggesting its potential for targeted screening even in low-prevalence settings.

### 3.4 Risk Stratification and Survival Phenotypes

#### 3.4.1 Unsupervised Phenotyping in the Development Cohort

The BGMM successfully identified three distinct risk strata (**Figure 3A & C**), with Kaplan-Meier analysis revealing a sharp divergence in outcomes (Log-rank p < 0.001). In both cross-validation and testing sets, the ‘High Risk’ group demonstrated an accelerated decline, indicating that the model successfully captures distinct survival phenotypes.

**Figure 3.**
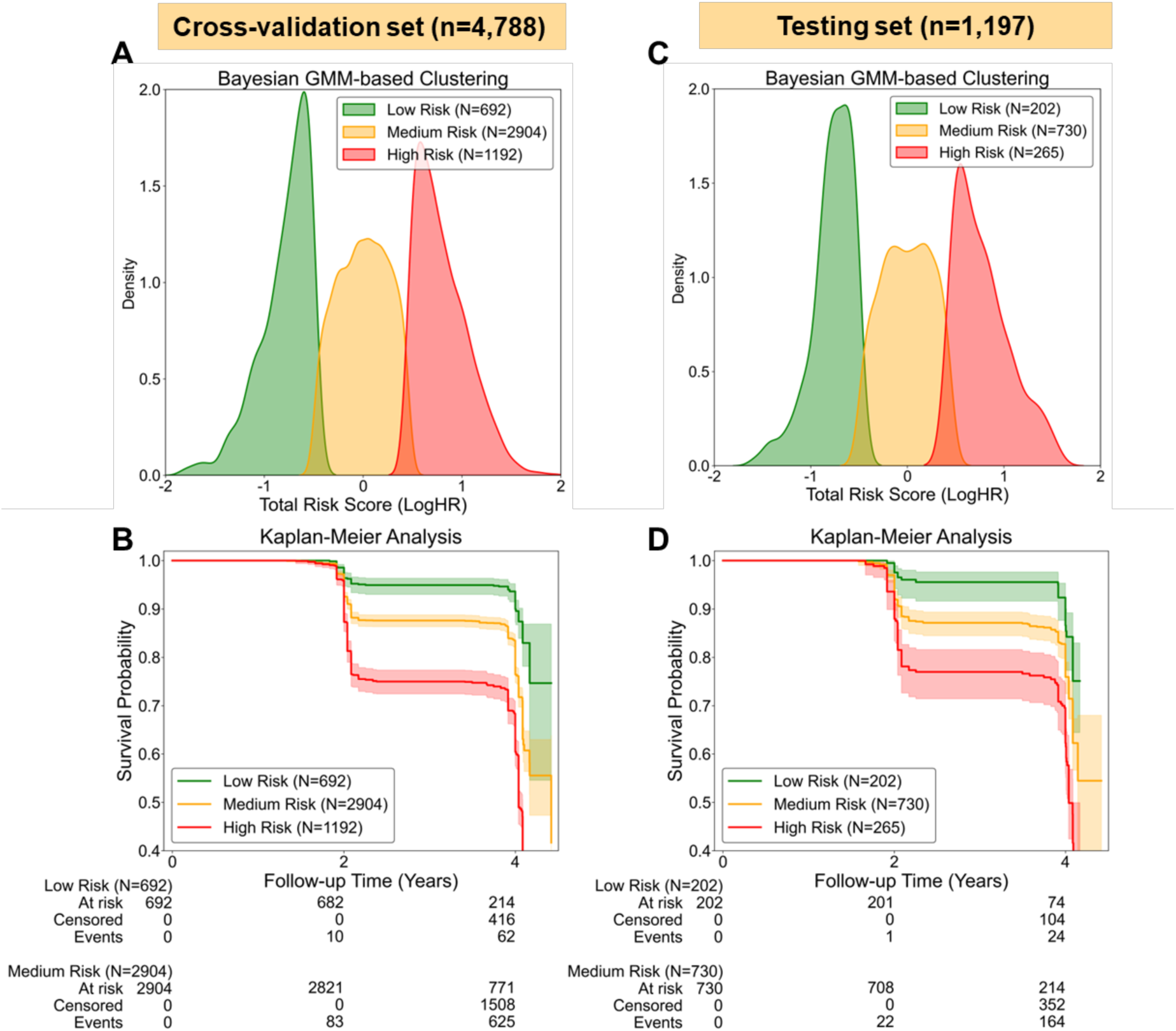
Risk Stratification Performance in Cross-validation and Internal Testing Sets. **(A,C)** Distribution of total risk scores (LogHR) derived from the CoxPH model. Bayesian GMM-based clustering identified three distinct risk phenotypes: Low Risk (green), Medium Risk (yellow), and High Risk (red). **(B, D)** Kaplan-Meier survival curves demonstrating significant separation in cognitive decline-free survival among the three identified risk groups (p < 0.001). The risk stratification logic remains consistent across both the 80% cross-validation set and the 20% internal testing set.

#### 3.4.2 Cross-cohort Risk Stratification

While the development cohort allowed for continuous longitudinal modeling, the validation cohorts were stratified via optimized decision thresholds (MaxStat). Despite the shorter follow-up, a clear gradient in cumulative incidence was observed (**Figure 4**). In the CHARLS validation, the 3-year incidence rose from 6.3% (Low Risk) to 38.0% (High Risk). Hence, these findings confirm the model’s ability to maintain a robust risk gradient across different populations.

**Figure 4.**
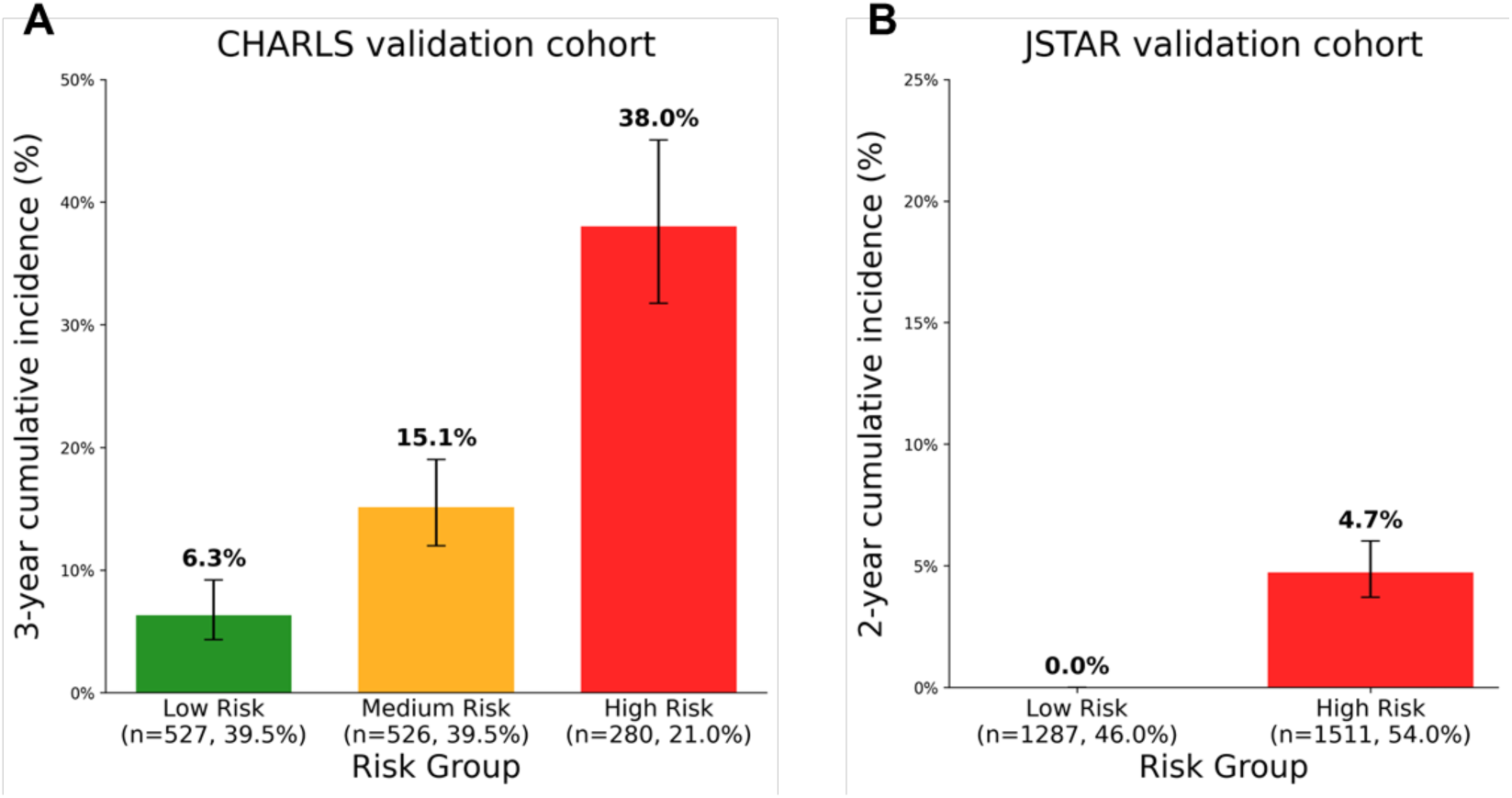
Risk stratification by predicted cognitive decline risk. Cumulative incidence of cognitive decline by model-defined risk groups in the validation cohorts: **(A)** CHARLS 2015-2018 and **(B)** JSTAR 2007-2009. (Cutoffs were pre-defined in the training set using MaxStat.)

### 3.5 Sensitivity Analyses

The results demonstrated remarkable temporal stability, as the 2-year CD-AUC (0.614) was nearly identical to the 4-year AUC (0.616) (**Supplementary Figure 4A**). This consistency suggests that the model captures robust risk features present early in the disease process. Furthermore, the 2-year zoomed calibration plot (**Supplementary Figure 4B**) showed excellent agreement in the lower risk deciles, justifying the model’s application to cohorts with shorter observation windows.

The complete-case analysis yielded performance metrics in the JSTAR cohort nearly identical to the primary analysis. Re-calibrating the preprocessor’s parameters to the JSTAR cohort while retaining the original Cox coefficients also resulted in similar performance (**Supplementary Table**). These findings suggest that the model’s validation performance is robust and not unduly influenced by the multiple imputation strategy.

## 4. Discussion

In the present study, a parsimonious risk predicting model with six predictors was developed in a large, representative Chinese cohort. The CoxPH model demonstrated moderate and robust performance in temporal validation within the same population. The model also exhibited consistent generalizability and transportability when validated in the external Japanese cohort, despite the geographical and temporal variations. To our knowledge, this is the first study to formally validate a cognitive decline prediction model developed in a Chinese cohort on a Japanese cohort.

Among all the linear and non-linear algorithms compared, the standard CoxPH model was identified as the optimal choice. This is not surprising, as the CoxPH model is a reliable and standard method for survival analysis. (25) On the other hand, its ability to surpass more complex ML algorithms suggests that the risk prediction for this outcome is adequately captured by a linear combination of these strong predictors. Nevertheless, GBSA, as the best-performing tree-based ensemble model, is also promising.

A key finding was observed when comparing internal and external validation performance. The model’s discrimination in the 20% internal hold-out test set was modest. In contrast, performance on the temporal and external validation was substantially higher. We believe the substantial case-mix difference between cohorts explains the higher C-index/AUC but poorer calibration observed in external validation: when the distribution of predicted risks is more polarized and events are concentrated among a small subgroup, rank-based metrics can appear higher even if overall calibration is poorer.

The final model’s discrimination is comparable to other regression-based studies, which typically report AUCs in the 0.7 range.(8,26) We are aware that biomarkers, such as genetics (e.g., APOE) or imaging, were not included, as the study aimed to develop a model for community-based screening using easily accessible features. While these biomarkers would likely improve performance substantially,(9,27) our findings demonstrate that a simple 6-predictor CoxPH model is sufficient for achieving robust risk stratification.

Overall, while the external follow-up duration was shorter (2–3 years) than the development cohort (4 years), the model maintained high consistency in its discriminative ability. However, this high discrimination was coupled with a significant probabilistic bias. The calibration plots indicate a systematic underestimation of absolute risk in the Japanese population. We speculate that this is likely attributable to the drastically lower baseline incidence of cognitive decline in the JSTAR cohort compared to the CHARLS training set. Despite this miscalibration, the risk stratification remained clinically robust: the model effectively identified a ‘zero-incidence’ low-risk stratum. These results suggest that while the model serves as an excellent relative risk ranking tool, absolute probability recalibration is necessary when shifting from a high-prevalence to an ultra-low-prevalence geriatric population.

DCA provided a detailed validation of the model’s clinical utility in the low-prevalence JSTAR cohort. While the positive net benefit was confined to a relatively narrow range of threshold probabilities near the population prevalence, this remains clinically significant for early-stage screening. In such a low-risk population, the ‘Treat None’ strategy is a solid baseline; however, the model still demonstrated superiority within the low-threshold window, suggesting it is best suited as a conservative first-stage screening tool to exclude individuals with near-zero risk rather than a tool for aggressive intervention.

Nonetheless, the mismatch between discriminative power and clinical utility is likely attributable to several factors. First, significant baseline differences in event dynamics between the Chinese development and Japanese validation cohorts create a tough scenario for model transportability. Second, a critical factor is the mismatch in follow-up duration; while our sensitivity analysis confirms that the model’s ranking ability (AUC) remains stable across 2-year and 4-year periods, its ability to estimate exact risk probabilities is naturally limited when the prediction time is shorter. These factors explain why the model can still identify high-risk individuals effectively (high AUC) even when the absolute probability values are not perfectly aligned (miscalibration) (28).

To summarize, these findings highlight a key conclusion: while the model’s ranking ability remained consistent and even improved across cohorts, its absolute risk estimates were sensitive to population differences and follow-up durations. This suggests that while the identified predictors are reliable for identifying high-risk individuals, the model requires local recalibration to provide accurate probability values in populations with different baseline risks.

### Limitations

While this study yielded a robust model for Chinese older adults and tested its generalizability in a Japanese population, several limitations should be acknowledged. First, the feature selection process relied on RSF-based permutation importance, which may favor variables with non-linear effects. However, the fact that the linear Cox model ultimately outperformed the non-linear approaches suggests that the selected predictors retained strong linear associations with the outcome. Second, a key limitation is the temporal and geographical mismatch between cohorts. The model was trained on 4-year follow-up data but validated on 2 to 3-year outcomes in a population with a much lower baseline incidence of cognitive decline. This discrepancy likely contributed to the observed miscalibration, suggesting that while the model’s risk ranking is stable, the absolute probability estimates require local recalibration before clinical use. Third, the validation JSTAR cohort had a high proportion of missing cognitive follow-up data (approx. 40%), which was handled using the same imputation pipeline as the training set. Nevertheless, our sensitivity analysis yielded nearly identical performance metrics, suggesting the results were not unduly influenced by the imputation strategy. Finally, future studies may focus on building models using combined cohort data across populations or developing formal strategies for model transportability and recalibration.

## Conclusion

In conclusion, we developed and validated a parsimonious 6-predictor Cox model that effectively identifies individuals at high risk for cognitive decline across different East Asian populations. Our findings demonstrate that while the model’s ranking ability is robust and generalizable—enabling the reliable identification of high-risk phenotypes—the absolute risk estimates are sensitive to geographical and temporal variations. This performance decoupling highlights the necessity of rigorous external validation even within biologically similar populations. Practically, our model serves as a potent ‘rule-out’ tool for low-prevalence settings, though population-specific recalibration remains a prerequisite for accurate clinical prognosis in new geographic contexts. These results provide a scalable framework for early cognitive risk screening in aging societies.

## Data availability

The datasets analyzed in this study are publicly available through the Gateway to Global Aging Data (https://g2aging.org/health-and-retirement-studies/overview).

## Code availability

The analytical code used in this study will be made available on GitHub upon publication.

## Acknowledgements

We thank the participants and staff of the CHARLS and JSTAR studies. JSTAR was conducted by the Research Institute of Economy, Trade and Industry (RIETI), Hitotsubashi University, and the University of Tokyo.

## Contributions

L.T. conceived and designed the study, performed the statistical analyses, interpreted the data, and drafted the manuscript. M.C., Y.N. and Y.K. contributed to data interpretation and critical revision of the manuscript. D.G. contributed to machine learning analyses and data interpretation. All authors read and approved the final manuscript. L.T. had primary responsibility for the final content.

## Correspondence

Correspondence and requests for materials should be addressed to L.T.

---

**Supplemental Table.** Performance of the Final Model in Validation Cohorts.

| Model | C-index | CD-AUC | IBS | RMSE |
| --- | --- | --- | --- | --- |
| CoxPH Model without elastic net penalty – Hold-out Validation | 0.609 (0.566–0.651) | 0.645 (0.606–0.685) | 0.114 (0.103–0.128) | **0.007** |
| Final model – Temporal Validation (CHARLS) | **0.722 (0.677–0.764)** | **0.701 (0.641–0.756)** | 0.066 (0.059–0.074) | 0.072 |
| Final model – External Validation (JSTAR) | 0.682 (0.654–0.713) | 0.689 (0.660–0.724) | **0.032 (0.028–0.037)** | 0.013 |
| **Sensitivity Analyses of External Validation (JSTAR)** |  |  |  |  |
| Complete-case Analysis | 0.678 (0.638–0.721) | 0.693 (0.646–0.735) | 0.028 (0.023–0.036) | 0.017 |
| Distributional Shift Test | 0.681 (0.681–0.682) | 0.688 (0.688–0.689) | 0.032 (0.032–0.032) | 0.013 |

Values represent the mean and 95% confidence interval.
C-index, concordance index based on inverse probability of censoring weights; CD-AUC, cumulative dynamic area under the curve; IBS, integrated Brier score; RMSE, root mean square error.

**Supplemental Figure 1.**
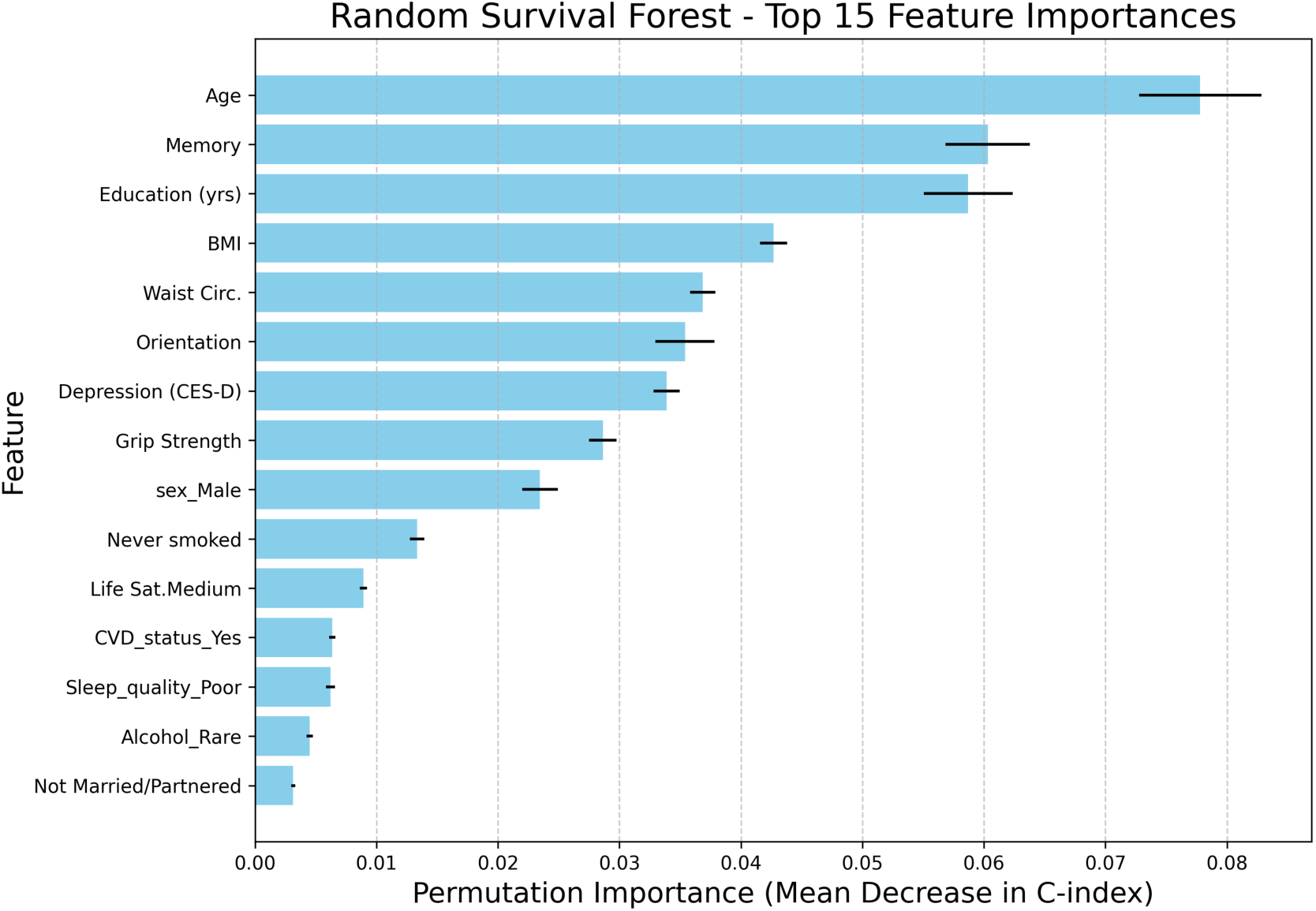
Permutation-based feature importance from the RSF model. Features are ranked by the mean decrease in the C-index across 10 repeats. Error bars represent the standard deviation of the importance metric.

**Supplementary Figure 2.**
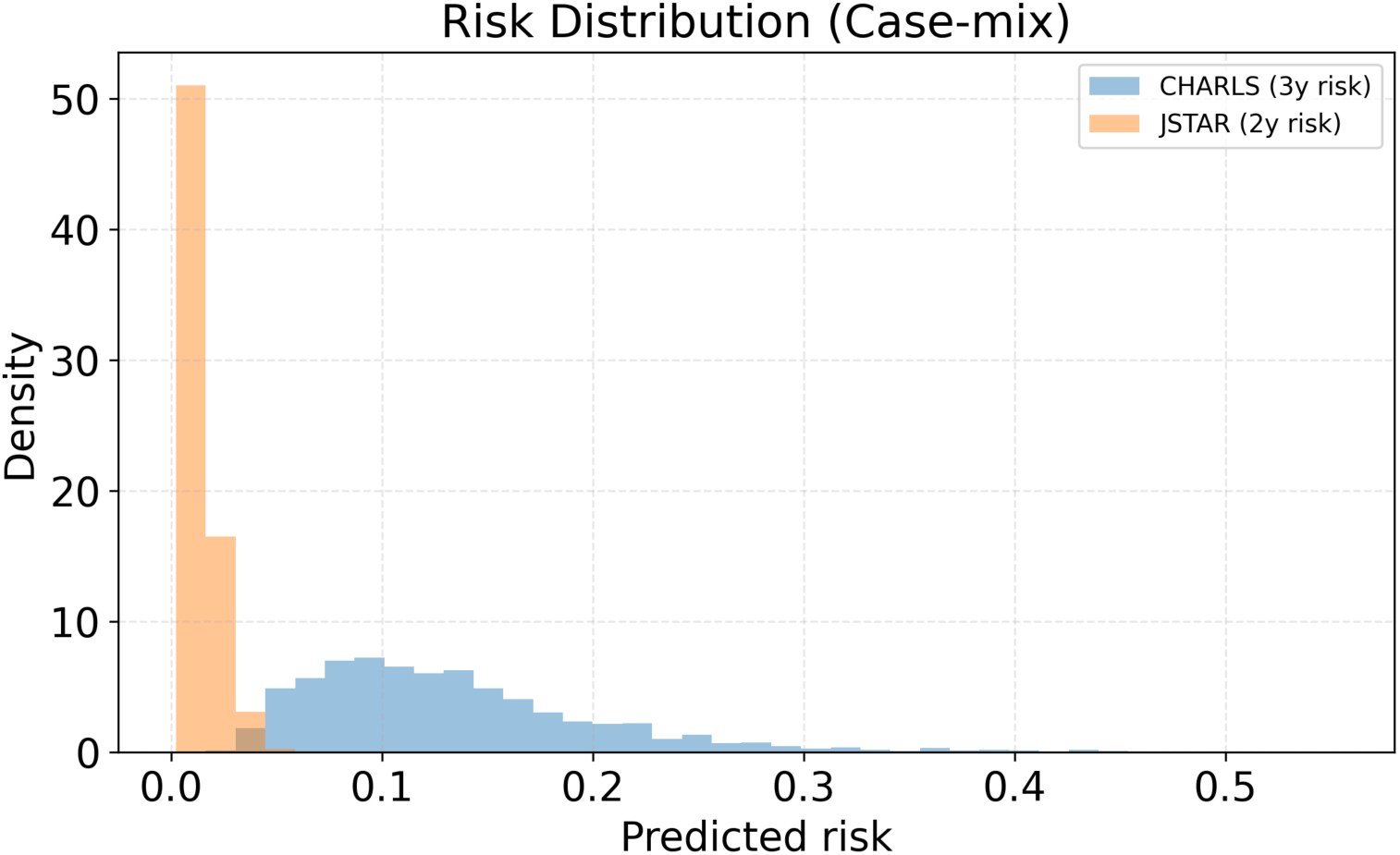
Predicted risk distribution across validation cohorts (case-mix comparison). Density histograms of model-predicted risks in the two validation cohorts. The CHARLS cohort (blue) exhibits a broad risk spread consistent with a moderate event rate, while the JSTAR cohort (orange) shows a highly skewed distribution with most predicted risks near zero, reflecting its very low observed incidence.

**Supplemental Figure 3.**
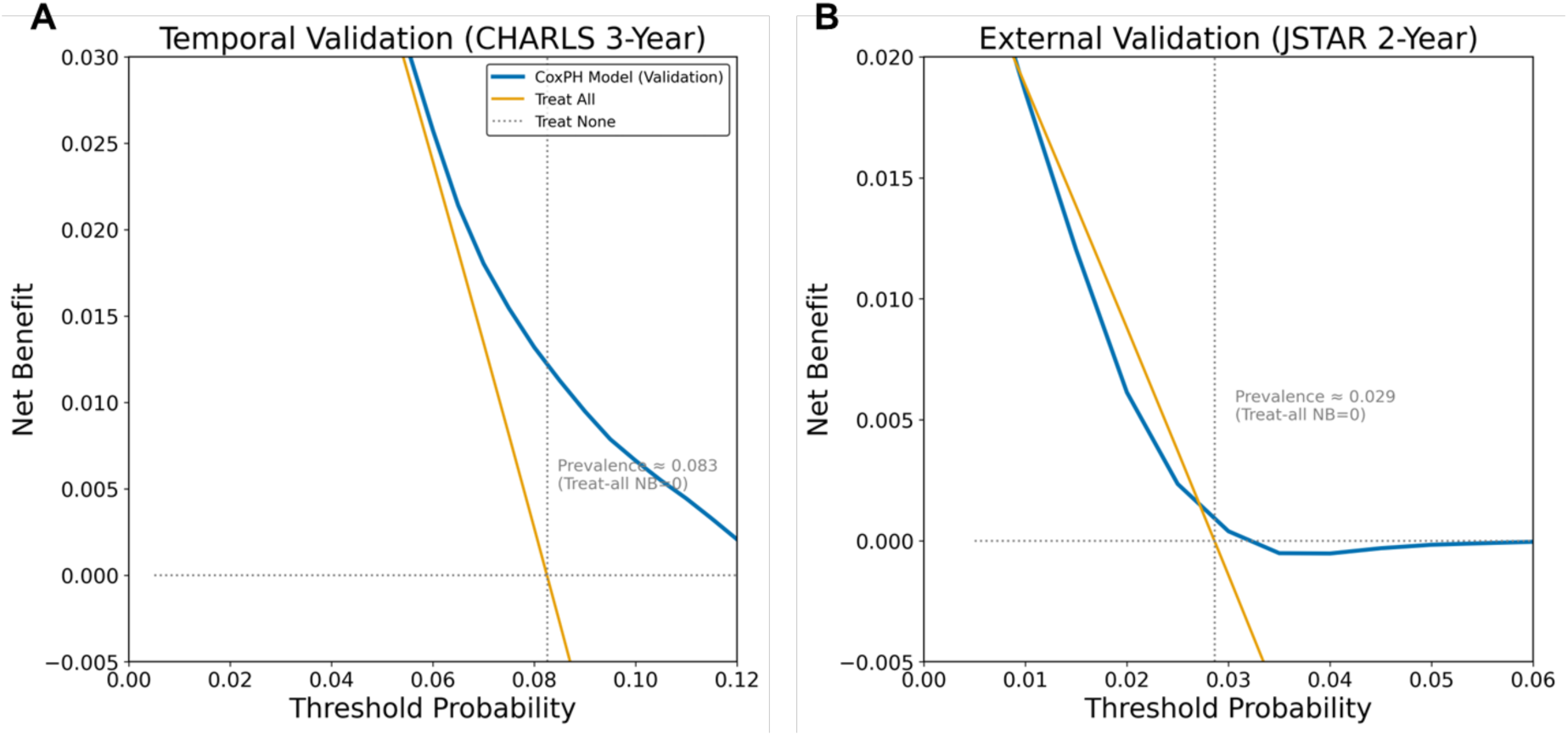
Clinical utility evaluated via Decision Curve Analysis (DCA) in validation cohorts. **(A)** In the temporal validation cohort (CHARLS 3-year), the CoxPH model demonstrated a consistent positive net benefit over both "treat-all" (orange line) and "treat-none" (dotted line) strategies across a clinically relevant threshold range of approximately 0.05 to 0.12. **(B)** In the external validation cohort (JSTAR 2-year), despite the extremely low prevalence (∼0.029), the model maintained a marginal but positive clinical utility at lower threshold probabilities. This suggests the model’s potential value in identifying high-risk individuals for targeted intervention even in low-incidence settings.

**Supplemental Figure 4.**
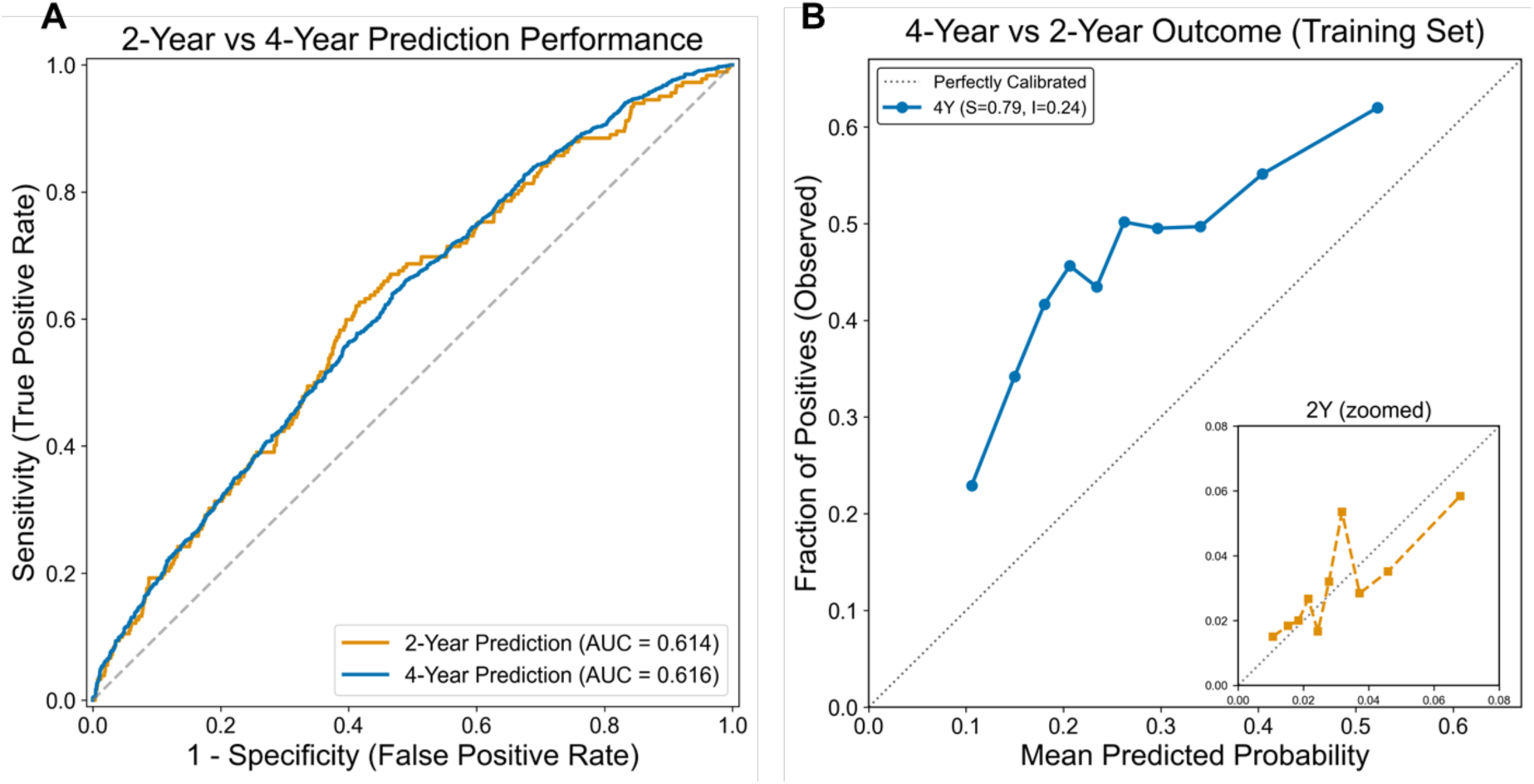
Sensitivity analysis of model performance at different follow-up horizons (Training Set). **(A)** ROC Analysis: Comparison of discriminative ability between 2-year and 4-year prediction horizons. The model achieved highly consistent results with an AUC of 0.614 at 2 years and 0.616 at 4 years. **(B)** Calibration Profiles: Calibration curves for 4-year (main plot) and 2-year (inset plot) outcomes. The 4-year calibration showed a slope of 0.79 and intercept of 0.24. The zoomed inset for the 2-year horizon demonstrates the model’s reliability in predicting early-stage cognitive decline events.

## References

1. 2025 Alzheimer’s disease facts and figures. Alzheimers Dement. 2025;21(4):e70235.

2. Jia L, Quan M, Fu Y, Zhao T, Li Y, Wei C, et al. Dementia in China: epidemiology, clinical management, and research advances. Lancet Neurol. 2020 Jan 1;19(1):81–92.

3. Zhang J, Zhang Y, Wang J, Xia Y, Zhang J, Chen L. Recent advances in Alzheimer’s disease: mechanisms, clinical trials and new drug development strategies. Signal Transduct Target Ther. 2024 Aug 23;9(1):211.

4. Livingston G, Huntley J, Liu KY, Costafreda SG, Selbæk G, Alladi S, et al. Dementia prevention, intervention, and care: 2024 report of the Lancet standing Commission. The Lancet [Internet]. 2024 Jul 31 [cited 2024 Aug 7];0(0). Available from: https://www.thelancet.com/journals/lancet/article/PIIS0140-6736(24)01296-0/fulltext

5. Tu L, Lv X, Yuan C, Zhang M, Fan Z, Xu X, et al. Trajectories of cognitive function and their determinants in older people: 12 years of follow-up in the Chinese Longitudinal Healthy Longevity Survey. Int Psychogeriatr. 2020 Apr 27;1–11.

6. Wu Z, Phyo AZZ, Al-harbi T, Woods RL, Ryan J. Distinct Cognitive Trajectories in Late Life and Associated Predictors and Outcomes: A Systematic Review. J Alzheimers Dis Rep. 2020 Dec 28;4(1):459–78.

7. Graham SA, Lee EE, Jeste DV, Van Patten R, Twamley EW, Nebeker C, et al. Artificial intelligence approaches to predicting and detecting cognitive decline in older adults: A conceptual review. Psychiatry Res. 2020 Feb;284:112732.

8. Sakal C, Li T, Li J, Li X. Identifying Predictive Risk Factors for Future Cognitive Impairment Among Chinese Older Adults: Longitudinal Prediction Study. JMIR Aging. 2024 Mar 22;7:e53240.

9. Tan WY, Hargreaves CA, Dawe GS, Hsu W, Lee ML, Vipin A, et al. Incremental Value of Multidomain Risk Factors for Dementia Prediction: A Machine Learning Approach. Am J Geriatr Psychiatry. 2025 Mar 1;33(3):229–44.

10. Qi W, Zhu X, Wang B, Shi Y, Dong C, Shen S, et al. Alzheimer’s disease digital biomarkers multidimensional landscape and AI model scoping review. Npj Digit Med. 2025 Jun 16;8(1):366.

11. You J, Zhang YR, Wang HF, Yang M, Feng JF, Yu JT, et al. Development of a novel dementia risk prediction model in the general population: A large, longitudinal, population-based machine-learning study. eClinicalMedicine [Internet]. 2022 Nov 1 [cited 2025 Mar 17];53. Available from: https://www.thelancet.com/journals/eclinm/article/PIIS2589-5370(22)00395-9/fulltext

12. D’Amore FM, Moscatelli M, Malvaso A, D’Antonio F, Rodini M, Panigutti M, et al. Explainable machine learning on clinical features to predict and differentiate Alzheimer’s progression by sex: Toward a clinician-tailored web interface. J Neurol Sci. 2025 Jan 15;468:123361.

13. Vermeulen RJ, Andersson V, Banken J, Hannink G, Govers TM, Rovers MM, et al. Limited generalizability and high risk of bias in multivariable models predicting conversion risk from mild cognitive impairment to dementia: A systematic review. Alzheimers Dement. 2025;21(4):e70069.

14. Lyall DM, Kormilitzin A, Lancaster C, Sousa J, Petermann-Rocha F, Buckley C, et al. Artificial intelligence for dementia—Applied models and digital health. Alzheimers Dement. 2023 Dec;19(12):5872–84.

15. Myszczynska MA, Ojamies PN, Lacoste AMB, Neil D, Saffari A, Mead R, et al. Applications of machine learning to diagnosis and treatment of neurodegenerative diseases. Nat Rev Neurol. 2020 Aug;16(8):440–56.

16. Ho SY, Phua K, Wong L, Goh WWB. Extensions of the External Validation for Checking Learned Model Interpretability and Generalizability. Patterns [Internet]. 2020 Nov 13 [cited 2025 Oct 7];1(8). Available from: https://www.cell.com/patterns/abstract/S2666-3899(20)30170-7

17. Azar SG, Tronchin L, Simkó A, Nyholm T, Löfstedt T. From Promise to Practice: A Study of Common Pitfalls Behind the Generalization Gap in Machine Learning. Trans Mach Learn Res [Internet]. 2024 Aug 27 [cited 2025 Oct 7]; Available from: https://openreview.net/forum?id=DqWvxSQ1TK

18. Collins GS, Dhiman P, Ma J, Schlussel MM, Archer L, Van Calster B, et al. Evaluation of clinical prediction models (part 1): from development to external validation. BMJ. 2024 Jan 8;384:e074819.

19. Zhao Y, Hu Y, Smith JP, Strauss J, Yang G. Cohort Profile: The China Health and Retirement Longitudinal Study (CHARLS). Int J Epidemiol. 2014 Feb 1;43(1):61–8.

20. Ichimura H, Shimizutani S, Hashimoto H. JSTAR First Results 2009 Report [Internet]. Research Institute of Economy, Trade and Industry (RIETI); 2009 Sep [cited 2025 Oct 7]. (Discussion papers). Available from: https://EconPapers.repec.org/RePEc:eti:dpaper:09047

21. Gauthier S, Reisberg B, Zaudig M, Petersen RC, Ritchie K, Broich K, et al. Mild cognitive impairment. The Lancet. 2006 Apr 15;367(9518):1262–70.

22. Bouzid Z, Sejdic E, Martin-Gill C, Faramand Z, Frisch S, Alrawashdeh M, et al. Electrocardiogram-based machine learning for risk stratification of patients with suspected acute coronary syndrome. Eur Heart J. 2025 Mar 7;46(10):943–54.

23. Du Y, Hu N, Yu Z, Liu X, Ma Y, Li J. Characteristics of the cognitive function transition and influencing factors among Chinese older people: An 8-year longitudinal study. J Affect Disord. 2023 Mar 1;324:433–9.

24. Kunutsor SK, Isiozor NM, Voutilainen A, Laukkanen JA. Handgrip strength and risk of cognitive outcomes: new prospective study and meta-analysis of 16 observational cohort studies. GeroScience. 2022 Aug 1;44(4):2007–24.

25. Yuan S, Liu Q, Huang X, Tan S, Bai Z, Yu J, et al. Development of an individualized dementia risk prediction model using deep learning survival analysis incorporating genetic and environmental factors. Alzheimers Res Ther. 2024 Dec 30;16(1):278.

26. Yuan C, Linn KA, Hubbard RA. Algorithmic Fairness of Machine Learning Models for Alzheimer Disease Progression. JAMA Netw Open. 2023 Nov 7;6(11):e2342203.

27. Shah J, Krell-Roesch J, Forzani E, Knopman DS, Jack CR, Petersen RC, et al. Predicting cognitive decline from neuropsychiatric symptoms and Alzheimer’s disease biomarkers: A machine learning approach to a population-based data. J Alzheimer’s Dis. 2025 Feb 1;103(3):833–43.

28. Efthimiou O, Seo M, Chalkou K, Debray T, Egger M, Salanti G. Developing clinical prediction models: a step-by-step guide. BMJ. 2024 Sep 3;386:e078276.

